# Some Clinical and Immunological Features of Imported COVID-19 Cases in Mongolia

**DOI:** 10.1101/2021.03.17.21253849

**Authors:** Munkh-Undrakh Batmunkh, Enkhsaikhan Lkhagvasuren, Oyungerel Ravjir, Tsogtsaikhan Sandag

## Abstract

SARS-CoV-2 disturbs the normal immune responses causing an uncontrolled inflammatory response in patients with severe COVID-19. The pattern of the immune response to the SARS-CoV-2 in individuals may fluctuate. Some have a virus-dependent protective immune response resulting in asymptomatic or mild disease with elimination of the virus within 7-10 days after onset of infection. Others develop virus non-dependent uncontrolled hyper-inflammation in the later period, leading to severe disease with cytokine storm, acute respiratory distress syndrome, disseminated intravascular coagulation and multi-organ failure.

**Methods:** The serum of 72 patients was investigated for titers of 15 cytokines and chemokines using Enzyme-linked immunosorbent assay (ELISA) kits in the serum of peripheral blood samples. The means of groups were compared using ANOVA followed by Tukey multiple post hoc comparisons if the ANOVA p-value was <0.05.

**Results:** Patients with pulmonary infiltrates on CT demonstrated a lower percentage of eosinophils (1.38±1.46%) and elevated level of serum CRP (8.57±19.10 mg/dL) compared to patients without pulmonary infiltrates (2.52±1.47% and 1.96±3.02 mg/dL respectively; p<0.05). ROC analysis for patients aged ≥35 years showed patients with mild disease (n=3) had a significantly higher titer of IL-1β and MCP-1 (AUC, 0.958 and 0.917 respectively, p<0.05) compared to patients with moderate disease (n=7).

## Introduction

Mongolia was recognized as a country without local transmission of the SARS-CoV-2 virus for about ten months because of its early and complex response to the pandemic(1). Local spread within Mongolia was prevented until November11, 2020, when its first case of local transmission case was identified, followed by clusters of other cases. According to a report of the Ministry of Health of Mongolia from December 31, 2020, there were 1215 confirmed cases of COVID-19 in the country, including 375 active and 830 recovered cases, and 399 of total cases was identified as imported (2).

It has been shown that SARS-CoV-2 disrupts the normal immune responses leading to an uncontrolled inflammatory response in patients with severe COVID-19(3). The pattern of the immune response to the SARS-CoV-2 in individuals may vary widely. Some have a virus-dependent protective immune response resulting in asymptomatic or mild disease with elimination of the virus within 7-10 days after onset of infection. Others develop virus non-dependent uncontrolled hyper-inflammation in the later period, leading to severe disease with cytokine storm, acute respiratory distress syndrome, disseminated intravascular coagulation and multi-organ failure (3-5). A better understanding of these events could contribute to the design of differential therapeutic approaches, depending on the disease stage, and to the delineation of prognostic and predictive biomarkers. Unfortunately, there are very few studies on the immune response in infected asymptomatic individuals, which would allow a better characterization of the protective immune response as it occurs under the natural conditions of the infection process(4).

This study analyzed and compared some hematological and immunological characteristics of COVID-19 patients with asymptomatic, mild and moderate disease. All of these cases were imported to Mongolia before September of 2020 from Mongolian nationals’ repatriation from pandemic countries and regions.

## Materials and methods

### Study population

A total of 276 cases of COVID-19 were detected among Mongolian citizens who were repatriated to the home country from pandemic countries and regions before July 31, 2020. All repatriated subjects were immediately tested for SARS-CoV-2 using a nucleic acid amplification test (NAAT). Positive cases were hospitalized in the National Center for Communicable Diseases, the center designated by the Mongolia Ministry of Health with public health related to infectious disease, patient care and research functions relating to the control and management of infectious diseases. Negative cases were transferred to the special isolation facilities approved by the State Emergency Commission of Mongolia and were retested using NAAT after 5-7 days. The clinical severity of each patient was classified using the World Health Organization’s (WHO) guidance for Clinical Management of COVID-19 from May 27, 2020 (6).

### Serum collection

We studied the serum of 72 of the 276 patients from March 10 to July 31, 2020, retrospectively after their peripheral blood was collected for clinical care purposes. A patient’s peripheral blood serum was included if it had been collected within the first 24 hours of hospitalization, there was sufficient volume, and had been stored at −16°C. We included all moderate cases with suitable blood samples. Asymptomatic or mild cases were randomly sampled from observation patients.

### Cytokine studies

The serum of 72 patients was investigated for titers of 15 cytokines and chemokines using Enzyme-linked immunosorbent assay (ELISA) kits (Sunlong Biotech, China) in the serum of peripheral blood samples. The following cytokines and chemokines were measured: pro-inflammatory cytokines - interleukins (IL-1β, IL-2, IL-6, IL-8, IL-12, IL-17), interferon-gamma (IFN*γ*) and tumor necrosis factor alpha (TNFα); anti-inflammatory cytokines (IL-4 and IL-10); chemokines –CXCL1, CCL2 (or monocyte attractant protein 1, MCP1) and IP-10 (interferon-gamma inducible protein 10 or CXCL10); and growth factors – G-CSF (Granulocyte Colony Stimulating Factor) and GM-CSF (Granulocyte-Monocyte Colony Stimulating Factor).

### Anti-SARS-CoV-2 immunoglobulins

The presence of anti-S-IgM and anti-S-IgG antibodies was determined using chemiluminescence immunoassay (MAGLUMI 2019-nCoV IgM/IgG, Snibe Co. Ltd., Shenzhen, China) in the serum of 55 patients.

### Chart review

The demographic, epidemiological and clinical data of all patients, including their hematology and C-reactive protein lab results in their early hospitalization and chest computed tomography (CT) findings later during the hospitalization, were collected from their medical records.

### Statistical analyses

We analyzed our data using the methods of descriptive and analytical statistics. The means of groups were compared using ANOVA followed by Tukey multiple post hoc comparisons if the ANOVA p-value was <0.05. The distribution of nominal variables among the patient groups was compared using Pearson’s chi-square test and Fisher’s exact test. The sensitivity and diagnostic ability of hematological differential and cytokine variables to classify disease were evaluated using receiver operating characteristic (ROC) analysis. Statistical calculations were performed using SPSS-17 software.

## Results

### Patients

SARS-CoV-2 was detected in 58 (80.5%) patients repatriated from Russia, 11 from Afghanistan, 3 from Kazakhstan, and 2 and 1 from Sweden and the USA, respectively. A total of 20 patients (27.8%) were classified as asymptomatic and 52 patients (72.2%) as symptomatic. Over two-thirds of hospitalized patients were diagnosed with a mild clinical form of the COVID-19 (190/276, 68.8%), and the remaining patients had a moderate form of the disease (86/276, 31.2%). None of the patients had severe disease, and there were no virus-related fatalities. Pulmonary infiltrates by chest CT were found in 15 patients with moderate disease later during their hospitalization. The demographic and epidemiological characteristics of patients according to clinical severity are shown in Table 1.

**Table 1.** Demographic, epidemiological, and clinical severity characteristics of 72 patients with COVID-19 repatriated to Mongolia from March to July 2020.

| Characteristics | Asympto-<br>matic | Symptomatic |  |  | Total<sup>-</sup> |
| --- | --- | --- | --- | --- | --- |
|  |  | Mild | Moderate | Subtotal |  |
| Age (years) |  |  |  |  |  |
| Mean (M ± SD)<sup>-</sup> | 24.7 ± 10.2 | 25.5 ± 10.0 | 29.6 ± 13.6 | 27.7 ± 12.1 | 26.9 ± 11.7 |
| Median | 21.0 | 21.5 | 23.5 | 22.0 | 22.0 |
| Max. – min. | 19 - 63 | 19 - 64 | 18 - 66 | 18 - 66 | 18 - 66 |
| Age groups, years, n (%)<sup>‡</sup> |  |  |  |  |  |
| 15-24 | 15 (20.8) | 19 (26.4) | 15 (20.8) | 33 (45.8) | 49 (68.1) |
| 25-34 | 2 (2.8) | 2 (2.8) | 6 (8.3) | 8 (11.1) | 10 (13.9) |
| 35-44 | 1 (1.4) | 3 (4.2) | 2 (2.8) | 5 (6.9) | 6 (8.3) |
| 45-54 | 0 | 0 | 3 (4.2) | 3 (4.2) | 3 (4.2) |
| 55-64 | 1 (1.4) | 1 (1.4) | 1 (1.4) | 2 (2.8) | 3 (4.2) |
| >65 | 0 | 0 | 1 (1.4) | 1 (1.4) | 1 (1.4) |
| Total | 19 (26.4) | 25 (34.7) | 28 (38.9) | 52 (72.2) | 72 (100.0) |
| Gender, n (%)<sup>‡</sup> |  |  |  |  |  |
| Males | 15 (20.8) | 15 (20.8) | 18 (25.0) | 33 (45.8) | 48 (66.7) |
| Females | 4 (5.6) | 10 (13.9) | 10 (13.9) | 19 (26.4) | 24/33.3 |
| Total | 19 (26.4) | 25 (34.7) | 28/38.9 | 52/72.2 | 72/100.0 |
| Lung involvement<sup>§</sup>, n (%)<sup>‡</sup> |  |  |  |  |  |
| Focal pneumonia | 0 | 0 | 15/20.8 | 15/20.8 | 15/20.8 |
| No pneumonia | 19 (26.4) | 25 (34.7) | 13/18.1 | 37/51.4 | 57/79.2 |
| Total | 19 (26.4) | 25 (34.7) | 28/38.9 | 52/72.2 | 72/100.0 |

□-total of asymptomatic and symptomatic
□-mean value and its standard deviation;
‡-observed cases (percent from total cases)
§-pulmonary infiltrate was found by CT

The most frequent symptoms observed in symptomatic patients were headache 14 (26.9%), dry cough (14 (26.9%), loss of smell (14(26.9%), nasal congestion (14/26.9%) and fever (10/19.2%) and taste loss (7(13.5%). Notably, most of our patients were young (median age 22 years), largely related to the fact that the majority or 55 of 72 (76.4%) were undergraduate university students repatriated from Russia. The mean age in moderate COVID-19 patients and distribution of moderate cases tended to be higher among older patients; however, the difference was not significant. In 15 (57.7%) of 26 moderate cases, focal infiltrates in lung parenchyma were found later during their hospitalization.

The hematological and blood chemistry features of patients determined in the early period of observation are shown in Table 2.

**Table 2.** Hematological profiles of COVID-19 measured within 24 hours of hospitalization stratified by disease severity (n=72).

| Blood cells | Asymptomatic | Symptomatic |  |  | Total<sup>-</sup> |
| --- | --- | --- | --- | --- | --- |
|  |  | Mild | Moderate | Subtotal |  |
| WBC<sup>-</sup> (×10<sup>9</sup>/L) | 6.5 ± 1.6 | 5.9 ± 1.4 | 6.3 ± 1.6 | 6.2 ± 1.7 | 6.2 ± 1.7 |
| RBC‡ (×10<sup>12</sup>/L) | 4.8 ± 0.4 | 4.6 ± 0.6 | 4.8 ± 0.5 | 4.7 ± 0.5 | 4.7 ± 0.5 |
| Hemoglobin (g/L) | 14.5 ± 1.7 | 14.0 ± 1.9 | 14.4 ± 1.5 | 14.3 ± 1.7 | 14.3 ± 1.7 |
| Hematocrit (%) | 40.3 ± 4.4 | 37.1 ± 8.9 | 40.7 ± 3.8 | 39.0 ± 6.9 | 39.6 ± 6.3 |
| Neutrophils (%) | 51.3 ± 10.2 | 50.6 ± 14.2 | 55.6 ± 10.4 | 53.2 ± 12.5 | 52.7 ± 11.9 |
| Lymphocytes (%) | 36.5 ± 10.3 | 35.1 ± 11.1 | 32.2 ± 9.2 | 33.5 ± 10.1 | 34.3 ± 10.1 |
| Monocytes (%) | 8.3 ± 2.0 | 9.4 ± 2.8 | 9.4 ± 2.6 | 9.4 ± 2.7 | 9.1 ± 2.5 |
| Eosinophils (%) | 2.45 ± 1.13 | 2.71 ± 1.72 | 1.71 ± 1.51 | 2.22 ± 1.68 | 2.29 ± 1.53 |
| Basophils (%) | 0.49 ± 0.28 | 0.38 ± 0.21 | 0.36 ± 0.16 | 0.37 ± 0.18 | 0.40 ± 0.23 |
| NLR (per)<sup>#</sup> | 1.53 ± 0.79 | 1.76 ± 0.94 | 1.95 ± 1.22 | 1.86 ± 1.09 | 1.77 ± 1.02 |
| Platelet (×10<sup>9</sup>/L) | 281.3 ± 54.0 | 275.5 ± 65.5 | 260.1 ± 63.3 | 267.4 ± 64.2 | 271.1 ± 61.6 |
| CRP (mg/dL) | 2.60±4.62 | 1.39±1.34 | 5.31±13.70 | 3.44±10.04 | 3.20±8.82 |

All values are mean ± standard deviation
□-total of asymptomatic and symptomatic
-White blood cells
‡Red blood cells
<sup>s</sup>neutrophil-to-lymphocyte ratio (in percent)

We did not find any significant differences in hematological and CRP values based on the patients’ clinical severity when comparing patients regardless of age. However, some differences were found to be age-dependent. When patients aged ≥35 years were analyzed separately, their WBC and CRP were accurate predictive classifiers for moderate versus mild disease (Figure 1).

**Figure 1.**
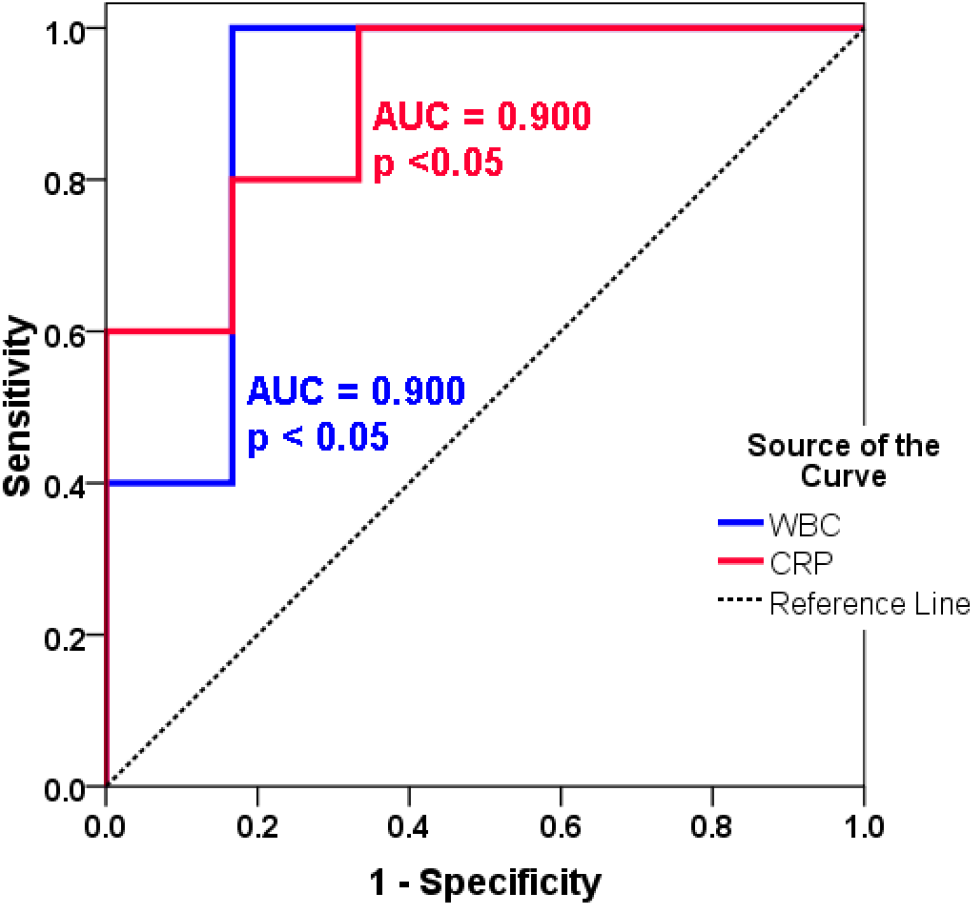
ROC analysis to detect the difference between mild and moderate disease in COVID-19 patients ≥ 35 years of age using white blood cell count (WBC) and C-reactive protein (CRP). Seven patients with moderate disease and four patients with mild disease were used in the analysis.AUC, Area under the Curve

Patients with pulmonary infiltrates on CT demonstrated a lower percentage of eosinophils (1.38±1.46%) and elevated level of serum CRP (8.57±19.10 mg/dL) compared to patients without pulmonary infiltrates (2.52±1.47% and 1.96±3.02 mg/dL respectively; p<0.05). However, only the eosinophil percentage was shown by itself as an accurate classifier for developing pulmonary infiltrates (Figure 2). The development of pulmonary infiltrates was not age-dependent.

**Figure 2.**
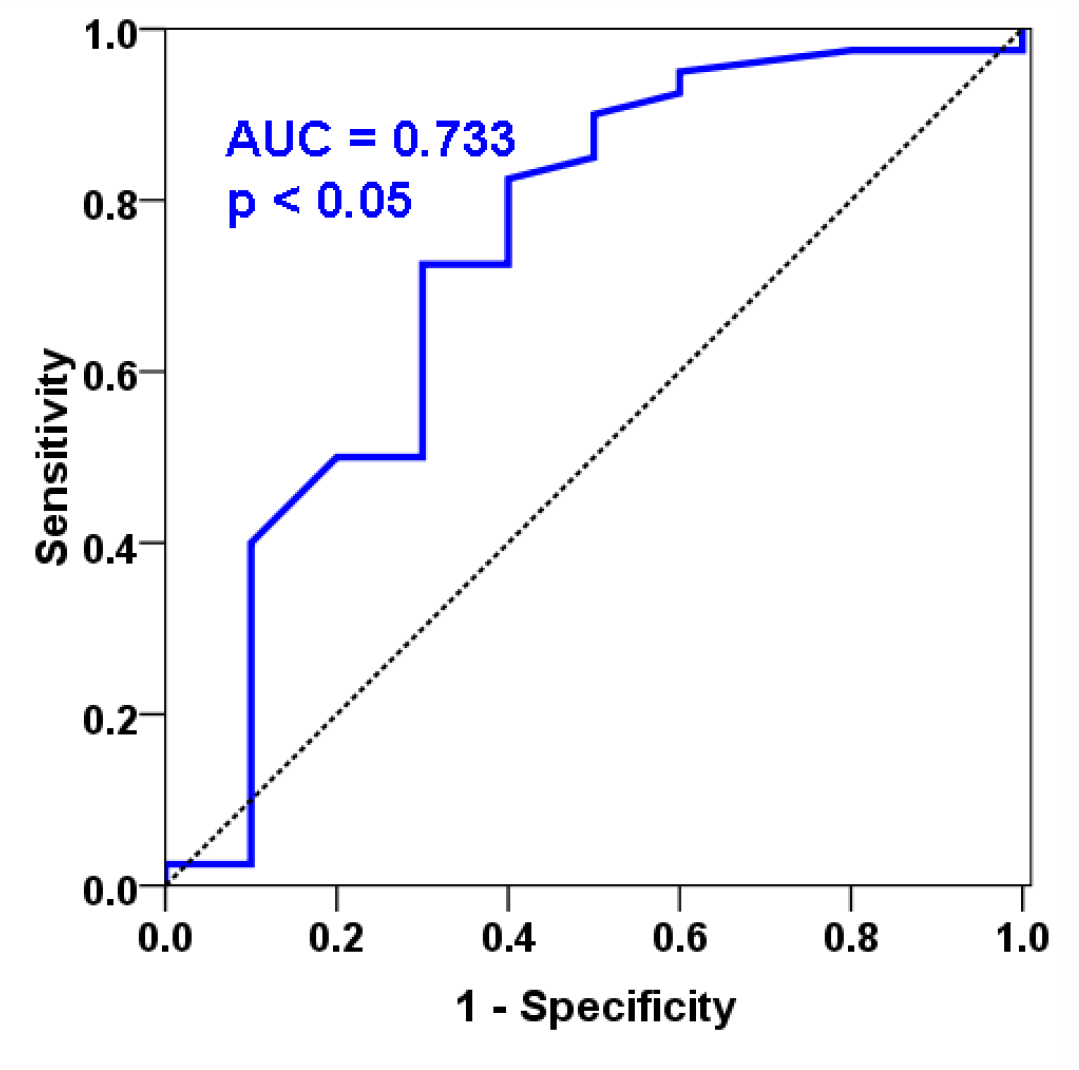
ROC analysis for using the eosinophil percentage to classify the subsequent development or absence of pulmonary infiltrates in COVID-19patientswith moderate disease regardless of age. Fifteen patients with pulmonary infiltrates and fifty-seven patients without infiltrates were used in the analysis. AUC, Area Under the Curve

The distribution of mean titers of serum cytokines and chemokines by the disease severity are shown in Table 4.

**Table 4.**
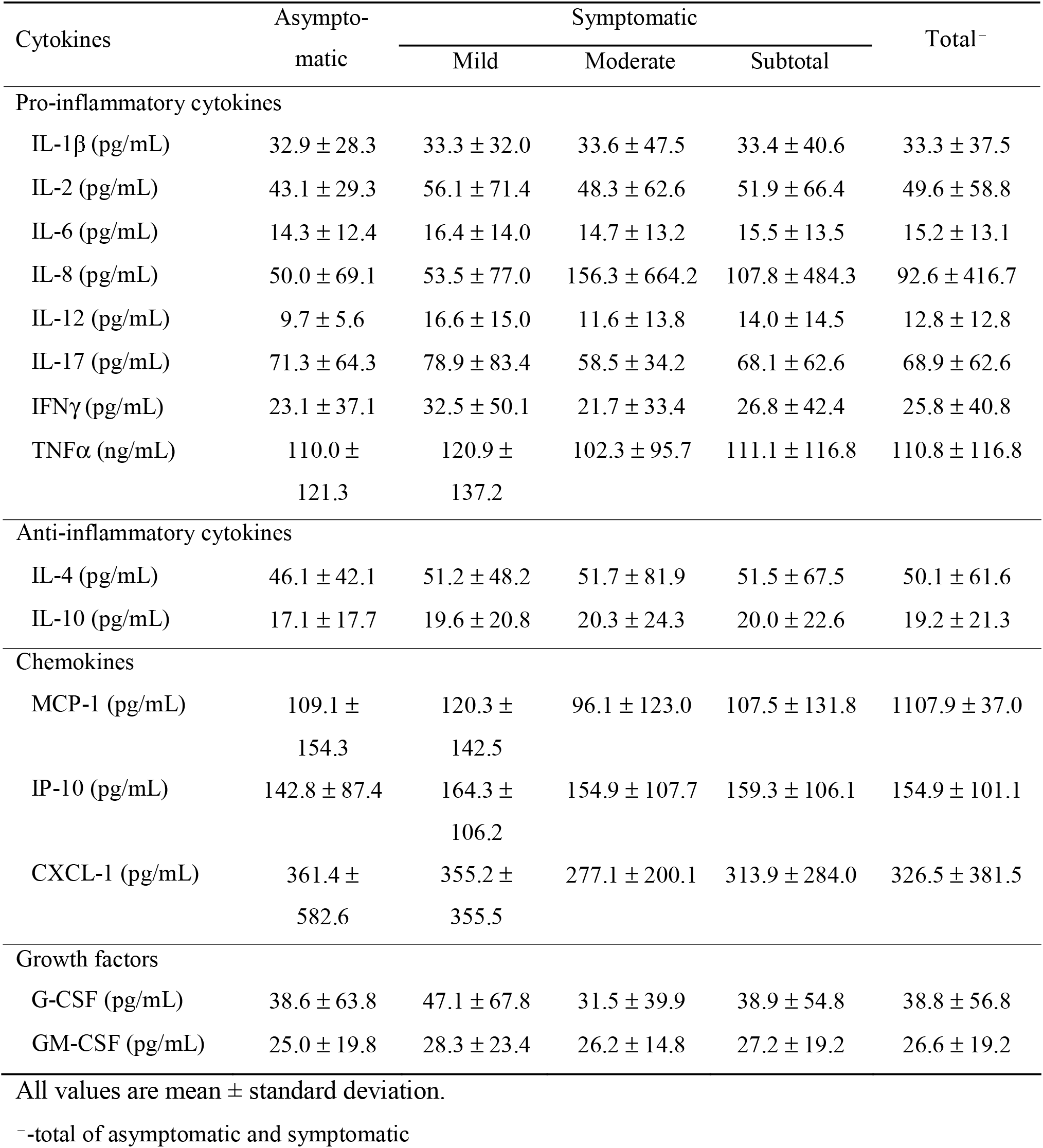
Serum cytokine and chemokines titers of COVID-19 patients measured within 24 hours of hospitalization stratified by disease severity (n=72).

None of the cytokine and chemokines group mean titers shown were significantly different. Furthermore, when was performed ROC analysis for patients aged ≥35 years, patients with mild disease (n=3) had a significantly higher titer of IL-1β and MCP-1 (AUC, 0.958 and 0.917 respectively, p <0.05) compared to patients with moderate disease (n=7).

Antibodies to SARS-CoV-2 (anti-S-IgM and anti-S-IgG) were investigated in 35 patients. Testing was performed within the first week of hospitalization for asymptomatic patients (6.5 ± 4.6 days) or after the onset of symptoms in symptomatic patients (6.6 ± 4.4 days). Anti-S-IgM was positive in 5 (29.4%) of 17 asymptomatic patients and 17 of 38 (44.7%) symptomatic patients (8 of 20 mild patients and 9 of 18 moderate patients). Anti-S-IgG was found in 13 (76.5%) of 17 asymptomatic patients and in 26 (68.4%) of 38 symptomatic patients (14 of 20 mild patients and 12 of 18 moderate patients). Anti-S-IgM were found in 4 of 7 patients (57.1%) and anti-S-IgG in 5 of 7 (71.4%) patients who developed pulmonary infiltrate, while anti-S-IgM were found in 37.5% and anti-S-IgG in 70.8% of 48 patients without lung involvement. The difference in the distribution of positive tests for anti-S-IgM and anti-S-IgG antibodies by disease severity and pulmonary injury was not significant (p>0.05).

## Discussion

Our study of patients with mild to moderate COVID-19 found few significant changes in peripheral blood and sera. This is probably because the majority of our patients were young and without chronic comorbidities and there were no cases of severe illness. However, when analyzing patients ≥35 years of age separately, the WBC and CRP were elevated in patients with moderate disease compared to patients with mild disease. In cases of severe infection, other authors have found the situation is somewhat different. For instance, Sun et al.(7) suggest that lower white blood cell counts, lower myeloid white blood cell counts, lower granulocyte counts, and higher eosinophil percentage increase the risk of severe COVID-19. Similar to our findings, another study demonstrated that higher CRP levels could be used as an independent factor to predict the severity of COVID-19(8). Mueller et al. suggested the increasing CRP levels during the first 48 hours of hospitalization are a better predictor (due to higher sensitivity) of respiratory decline than initial CRP levels (9). Li et al. found elevated CRP levels in COVID-19 patients compared to healthy controls and recommended using elevated CRP levels together with eosinopenia for differentiation of suspected COVID cases in an epidemic region having a large number of patients with COVID-19 and other respiratory diseases with resource-limited areas without available nucleic acid tests and radiographic examinations(10).

In our study, higher titers of IL-1β and MCP-1 were found in patients with mild disease ≥35 years of age than patients with moderate disease of the same age. There was only one patient >35 years with asymptomatic infection, and his titers of cytokines were comparable with those of moderate patients. Noroozi et al. reported very few sources of wide-spectrum analysis of cytokine profiles in patients with coronavirus infections (11). Ng et al. found increased IL-1β levels in children with severe acute respiratory syndrome (SARS) before the initiation of steroid therapy for severe disease (12).Min et al. showed increased MCP-1 levels in the plasma of patients with Middle East respiratory syndrome(MERS), but the cytokine titers were not associated with disease severity (13).

In our opinion, the most practical finding of our study was the reduced eosinophil percentage at hospital admission found in patients who later developed pulmonary infiltrates. The association of reduced peripheral blood eosinophil percentage, especially early in the course of the infection, with the severity and manifestation of hyperinflammatory response in the lung parenchyma has been reported in several studies (10, 14-17). We hope that our findings will help predict the severity of COVID-19 in medical and public health facilities.

### Study limitation

The authors suggested that relatively young age of patients and absence of cases with severe illness were restricted the chance to find enough significant differences in hematological and immunological features according to disease course of SARS-CoV-2 infected patients in this study.

## Data Availability

All data referred to in the manuscript are availiable as SPSS .sav files and can be provided on reqeust

## References

1. Erkhembayar R, Dickinson E, Badarch D, Narula I, Warburton D, et al. Early policy actions and emergency response to the COVID-19 pandemic in Mongolia: experiences and challenges. The Lancet. 2020;8(9):E1234-E41. DOI:https://doi.org/10.016/S2214-109X(20)30295-3.

2. Informed Consent Form Template for Clinical Studies. Research Ethics Review Committee (WHO ERC). https://www.who.int/ethics/review-committee/informed_consent/en/ [Internet]. 2018.

3. Yang L, Liu S, Liu J, Zhang Z, Wan X, et al. COVID-19: immunopathogenesis and Immunotherapeutics. Sig Transduct Target Ther. July 2020;5(128):doi.org/10.1038/s41392-020-00243-2.

4. Garcia LF. Immune Response, Inflammation, and the Clinical Spectrum of COVID-19. Front Immunol. June 2020;11(Article 1441):doi: 10.3389/fimmu.2020.01441.

5. Mortaz E, Tabarsi P, Varahram M, Folkerts G, Adcock IM. The Immune Response and Immunopathology of COVID-19. Front Immunol. 26 Aug 2020;11(Article 2037):doi.org/10.3389/fimmu.2020.02037.

6. Clinical Management of COVID-19. WHO Interim Guidance. WHO reference number: WHO/2019-nCoV/clinical/2020.5 27 May 2020.

7. Sun Y, Zhou J, Ye K. White blood cells and severe COVID-19: a Mendelian randomization study. MedRxiv. Oct 2020:doi.org/10.1101/2020.10.14.20212993.

8. Liu F, Li L, Xu MD, Wu J, Luo D, et al. Prognostic value of interleukin-6, C-reactive protein, and procalcitonin in patients with COVID-19. Journal of Clinical Virology. Apr 2020;127:DOI: 10.1016/j.jcv.2020.104370.

9. Mueller AA, Tamura T, Crowley CP, DeGrado JR, Haider H, et al. Inflammatory Biomarker Trends Predict Respiratory Decline in COVID-19 Patients. Cell Reports Medicine. Nov 2020;1(8):doi.org/10.1016/j.xcrm.2020.100144.

10. Li Q, Ding X, Xia G, Chen H-G, Chen F, Geng Z, et al. Eosinopenia and elevated C-reactive protein facilitate triage of COVID-19 patients in fever clinic: a retrospective case-control study. E Clinical Medicine. May 2020;23:100375.

11. Noroozi R, Branicki W, Pyrc K, Labaj P, Pospeich E, et al. Altered cytokine levels and immune responses in patients with SARS-CoV-2 infection and related conditions. Cytokine. Sep 2020;133:doi: 10.1016/j.cyto.2020.155143.

12. Ng PC, Lam CW, Li AM, Wong CK, Cheng FW, Leung TF. Inflammatory cytokine profile in children with severe acute respiratory syndrome. Pediatrics. Jan 2004113(1):e7–e14.

13. Min CK, Cheon S, Ha NY, Sohn KM, Kim Y, et al. Comparative and kinetic analysis of viral shedding and immunological responses in MERS patients representing a broad spectrum of disease severity. Sci Rep. May 20166(1):1–12.

14. Lindsley AW, Schwartz JT, Rothenberg ME. Eosinophil responses during COVID-19 infections and coronavirus vaccination. Journal of Allergy and Clinical Immunology. Jul 2020146(1):1-7. doi.org/10.1016/j.jaci.2020.04.021.

15. Xie G, Ding F, Han L, Yin D, Lu H, Zhang M. The role of peripheral blood eosinophil counts in COVID-19 patients. Allergy. June 2020;00p1– 12. https://doi.org/0.1111/all.14465.

16. Fraisse M, Logre E, Mentec H, Cally R, Plantefeve G, Contou D. Eosinophilia in critically ill COVID-19 patients: a French monocenter retrospective study. Critical Care. Nov 2020;24:635. doi.org/10.1186/s13054-020-03361-z.

17. Gonzalez MM, Gonzalo SE, Lopez CI, Fernandez AF, Perez BJL, Monge MD, et al. The prognostic value of eosinophil recovery in COVID-19: a multicentre, retrospective cohort study on patients hospitalised in Spanish hospitals. medRxiv. Aug 2020;2020(20172874).

